# Relationship between topological efficiency of white matter structural connectome and plasma biomarkers across Alzheimer’s disease continuum

**DOI:** 10.1101/2023.07.21.23292999

**Authors:** Mingkai Zhang, Haojie Chen, Weijie Huang, Tengfei Guo, Guolin Ma, Ying Han, Ni Shu

## Abstract

**Introduction:** The associations between plasma AD biomarkers, brain network topology, and cognition across AD continuum remained unknown.

**Methods:** From the cohort of Sino Longitudinal Study of Cognitive Decline, we analyzed plasma biomarkers of 287 participants, including the levels of β-amyloid (Aβ), phosphorylated-tau181 (p-tau181), glial-fibrillary-acidic-protein (GFAP), and neurofilament-light-chain (NfL), and we assessed white matter network efficiency of 395 participants. Trend analyses evaluated the sensitivity of plasma markers and network efficiency with AD progression. Correlation and mediation analyses further explored the relationships among plasma markers, network efficiency, and cognition across AD continuum.

**Results:** Among the plasma markers, GFAP exhibited the highest sensitivity with AD progression, followed by NfL, p-tau18, and Aβ42/Aβ40. Local efficiency decreased in multiple brain regions and correlated with GFAP, NfL, and p-tau181. Network efficiency mediated the relationship between plasma markers and cognition.

**Discussion:** Our findings highlight the potential of network-plasma approaches for early detection and intervention of AD.

## 1 Background

Alzheimer’s disease (AD) is a progressive neurodegenerative disorder characterized by cognitive decline, memory loss, and the inability to perform daily activities[1]. As the most common form of dementia, AD accounts for an estimated 60-80% of dementia cases worldwide, and the number of individuals affected expected to nearly triple by 2050[2,3]. The current lack of a cure and the irreversible nature of AD highlights the critical importance of early diagnosis, particularly at the preclinical stage. While biomarker detection in cerebrospinal fluid (CSF) and positron emission tomography (PET) imaging has advanced preclinical diagnosis[4], there remains a pressing need to explore practical, noninvasive, and cost-effective methods for identifying preclinical AD features.

A growing body of evidence implies that the pathophysiologic process of AD begins 15–20 years prior to the onset of clinical symptoms, especially in biomarkers and neuroimaging[4,5]. With the rapid development of ultra-sensitive assays, it is now possible to measure trace levels of brain-specific proteins in the blood[6]. Plasma biomarkers are playing an increasingly important role in this area. Recent studies have showed that amyloid-β (Aβ) 42/40 and phosphorylated tau (p-tau) from plasma appear to be the best candidate markers during both symptomatic AD as well as during preclinical AD[7,8]. While blood levels of neurofilament light (NfL) and glial fibrillary acidic protein (GFAP) are abnormal in a range of neurodegenerative disorders, nonetheless, there is a growing body of research that explores the potential of these markers as biomarkers for AD[9,10]. A recent investigation has revealed that plasma levels of GFAP experience a noteworthy elevation during the preclinical stage of AD[11]. Moreover, plasma GFAP exhibits the most effective performance for monitoring longitudinal disease progression [12]. Despite the potential of plasma biomarkers as diagnostic tools in routine clinical practice, their diagnostic efficacy alone may still fall short of being sufficient.

With the development of brain imaging and network modeling techniques, human brain connectomics has provided valuable insights into understanding the dynamic neuronal network changes from a system level in various brain disorders[13]. As a disconnection syndrome, AD is characterized by both structural and functional network changes throughout its continuum[14,15]. With diffusion MRI, our previous studies have demonstrated decreased topological efficiency of white matter (WM) structural network in amnestic mild cognitive impairment (MCI) and subjective cognitive decline (SCD) patients[16], especially involving key regions of default mode network (DMN) and striatum network [17]. Structural network efficiency measures the brain’s infrastructure supporting inter-neuronal communication [13]. Recent investigations have unveiled a significant correlation between network efficiency, AD pathology, and relevant biomarkers[15]. Specifically, a noticeable decline in local efficiency with AD progression has been detected[18], with these changes closely associated with hallmark pathological features, including Aβ deposition and tau tangles[19,20].

The interplay between pathological molecules in plasma and cognitive decline during the progression of AD constitutes a complex and multifaceted topic. One possible mechanism by which GFAP influences cognitive function is through glial activation and neuroinflammation[21]. Simultaneously, plasma levels of NfL may impact cognitive function in AD through various pathways, including brain atrophy, neuronal damage, and glucose hypometabolism[9]. Notably, we emphasize the mediating role of brain imaging-based biomarkers in the relationship between cognitive function and plasma biomarkers in AD[22]. However, there is a paucity of comprehensive studies investigating the relationship between plasma biomarkers, neuronal networks, and cognition. Thus, the objectives of the present study are threefold: (1) to explore the dynamic changes in plasma biomarkers related to AD, specifically GFAP, across different stages of the AD continuum, (2) to investigate the dynamic alterations in brain network efficiency within AD-related regions, and (3) to assess the associations between brain network efficiency, plasma markers, and cognitive performance across AD continuum.

## 2 Methods

### 2.1 Participants

In this study, all participants were enrolled in the Sino Longitudinal Study of Cognitive Decline (SILCODE) cohort. SILCODE is an ongoing prospective multicenter AD cohort study in the Han population of mainland China[23]. A total of 474 participants were recruited, and all participants were divided into different datasets depending on research needs: including plasma dataset (complete data on AD-related plasma markers were available for each participant), imaging dataset (each participant had complete imaging data available for brain network analysis), and combined dataset (participants in the first two datasets who matched plasma and imaging time differences of not more than 80 days, see Supplementary Figure 1).

The diagnosis of normal controls (NC) was based on the exclusion of MCI and dementia[24,25], requiring a CDR score of 0, no overt affective disorder, and normal education-adjusted scores on the MMSE and memory subdomain. Diagnostic criteria for SCD rely on defining and characterizing subjective cognitive decline[26]. The diagnosis of MCI was made based on neuropsychological criteria[25]. In order to qualify for AD dementia, the entry criteria must meet the proposed criteria for probable AD-induced dementia[24]. Eventually, the plasma dataset (n=287) includes 121 NC, 107 SCD, 41 MCI, and 18 AD. The imaging dataset (n=395) includes 129 NC, 112 SCD, 17 MCI, and 7 AD. The combined dataset (n=55) includes 25 NC, 17 SCD, 8 MCI, and 5 AD.

All participants or their legal guardians provided written informed consent. All procedures were approved by the Medical Research Ethics Committee and Institutional Review Board of Xuanwu Hospital. Cohorts are registered with ClinicalTrials.gov (SILCODE: NCT02225964). For each participant, clinical data were collected, including age, gender, years of education, and apolipoprotein E (APOE) genotype. Neuropsychological assessments focus on global cognitive function: Mini-Mental State Examination (MMSE) and Montreal Cognitive Assessment-Basic Version (MoCA-B)[27].

### 2.2 Plasma biomarkers

Blood samples (2 ml venous blood) were taken between 7: 00 and 8: 00 in the morning after an overnight fast using EDTA tubes, and blood samples were collected in the following manner. Supernatants were collected as plasma after being centrifuged several times for 15 min at 4°C (speed: 2500 g/min). All plasma samples were stored at −80°C and immediately thawed on ice prior to assay. Plasma levels of β-amyloid, p-tau181, GFAP, and NFL were measured using the Single Molecule Array (Simoa™) platform (Quanterix Corporation, Billerica, MA, USA) following the manufacturer’s instructions. The Simoa assays were performed using the following commercially available kits: β-amyloid (1-42) (Aβ42) and β-amyloid (1-40) (Aβ40) - Simoa™ β-Amyloid 42 (1-42) and β-Amyloid 40 (1-40) Advantage Kits, phosphorylated tau at threonine 181 (ptau181) Simoa™ p-tau181 Advantage Kit, Glial Fibrillary Acidic Protein (GFAP) Simoa™ GFAP Discovery Kit and Neurofilament light chain (NfL) - Simoa™ NF-light® Advantage Kit. All assays were performed in duplicate, and the average concentration values were reported. Sample concentrations below the lower limit of quantification (LLOQ) were assigned a value of half the LLOQ.

### 2.3 Image acquisition and preprocessing

In the SILCODE cohort, MRI data were acquired using an integrated simultaneous 3.0 T TOF PET/MR (SIGNA PET/MR, GE Healthcare, Milwaukee, Wisconsin, USA) at the Xuanwu Hospital of Capital Medical University, Beijing, China. About SMRI, the parameters for T1-weighted 3D brain structural images are as follows: SPGR sequence, FOV=256×256mm^2^, matrix=256×256, slice thickness=1mm, gap=0, slice number=192, repetition time (TR)=6.9 ms, echo time (TE)=2.98 ms, inversion time (TI)=450 ms, flip angle=12°, voxel size=1×1×1mm^3^. The DTI data were obtained with a single-shot spin-echo diffusion-weighted echo planar imaging (EPI) sequence with the following parameters: FOV=224×224mm^2^, data matrix=112×112, slice thickness=2mm, gap=0, slice number=70, slice order=interleaved, TR=16500 ms, TE=95.6 ms, 30 gradient directions (b=1000s/mm^2^) and 5 b0 images, voxel size=2×2×2mm^3^.

The preprocessing of diffusion MRI data involved denoising, head motion correction, and eddy current correction. Specifically, we first denoised the raw diffusion data with the **dwidenoise** command from MRtrix3 software (https://mrtrix.readthedocs.io/)[28], which produced a denoised diffusion-weighted data file. The **fslroi** command was then used to extract a single volume from the denoised data, which was used as the reference image for head motion correction and brain extraction. The **bet2** command was applied to this reference image to obtain the brain mask. Finally, eddy current correction was performed using the **eddy_openmp** command with the denoised data, brain mask, and gradient information as inputs.

### 2.4 Brain structural network construction

To construct the individual structural network, we used **bet** command to extract the brain and **fast** command to segment it into different tissue types on the T1-weighted image with FSL software (https://fsl.fmrib.ox.ac.uk/) [29]. The WM segmentation in T1 space was transformed into diffusion space and used as a seed mask for further tractography. **Bedpostx** command was then performed on the preprocessed diffusion MRI data to generate distributions on diffusion parameters at each voxel, with three fibres modeled per voxel, burn-in period set to 3000, and deconvolution model with sticks. Deterministic tractography was performed using a **track** command from CAMINO software (http://camino.cs.ucl.ac.uk/)[30] with the “bedpostx_dyad” input model and various parameters, including nearest-neighbor interpolation, Fourth-order Runge-Kutta method as the tracking algorithm, and tracking step size set to 2mm. Compartments with a mean volume fraction below 0.1 were discarded, and tracking was terminated if curvature exceeded 45 degrees at each 5 mm interval, and fibers with a length below 20mm or above 250mm were removed. To define network node, the BNA template were applied and transformed into individual diffusion space to parcellate the brain into 246 regions (https://atlas.brainnetome.org/) [31]. Tractography and brain parcellation were combined to generate a fiber-number weighted connectivity matrix with **conmat** command, representing the individual WM structural network.

### 2.5 Network efficiency computation

Based on the brain structural network, we calculated three network efficiency measures with BCT toolbox (https://www.nitrc.org/projects/bct/)[32], including global efficiency, local efficiency, and generalized local efficiency. Each measure was calculated at both regional and network levels. Here are the detailed descriptions of network efficiency measures:

***<u>Global efficiency</u>*** reflects the integration of region *i*, which was calculated as

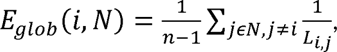

where the *L_i,j_* is the shortest path length between node *i* and node *j* within the network *N*.

***<u>Local efficiency</u>*** captures region *i*’s segregation and fault tolerance capabilities, which was computed as the mean of global efficiency across all nodes encompassed within *i*’s neighborhood:

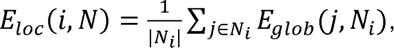

where *N_i_* is the sub-network of node *i* comprising *i*’s neighbors with the connections among them (Figure 3B).

***<u>Generalized local efficiency</u>*** enhances robustness against irrelevant factors and establishes weight-scale invariance[33], as calculated with

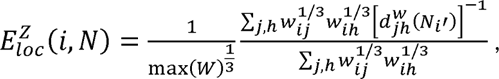

where *w_i,j_* is the weight (fibre-number) between region *i* and *j*, and 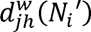 is the adapted shortest distance between region *j* and *h*, calculated as the shortest distance in the network *N_i_′* containing all neighbors of *i* excluding node *i* while considering a replacement of the weight of the edge (*j, h*) to 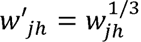.

### 2.6 Statistical analysis

The demographic variables were analyzed using ANOVA for continuous variables, including age, years of education, MMSE, MOCA, levels of plasma markers, and network efficiency. For discrete variables like sex, chi-squared tests were employed. These statistical analyses were performed across four diagnostic groups (NC, SCD, MCI, and AD) separately on three datasets: plasma, imaging, and combined datasets.

Trend analyses were performed on plasma markers, regional and whole-brain network efficiency, and cognitive performance among four diagnostic groups. For measures with significant linear or quadratic trends, *post-hoc* comparisons were performed with two sample t-tests. Plasma markers and network efficiency were analyzed separately within the plasma dataset and imaging dataset.

Correlation analyses were conducted among plasma markers, network efficiency, and cognitive performance across the AD continuum. Specifically, we evaluated the correlation between plasma markers and cognitive performances (MMSE and MOCA score) in the plasma dataset. In the combined set, we computed partial correlations between network efficiency, plasma markers, and cognitive performances controlling for covariates.

Furthermore, we tested the mediation of network efficiency on the relationship between plasma markers and cognition using the mediation package[34]. Bootstrapping procedures with 10,000 re-samplings were used to obtain the indirect effects’ significance and 95% confidence intervals.

All statistical analyses were performed using R (version 4.1.1). Covariates of age, sex, years of education and APOE4 status were removed in all the above analyses. On regional statistics, false discovery rate (FDR) corrections were applied in trend analyses and regional correlations. For *post-hoc* comparisons, Bonferroni corrections were applied. We considered significance at a nominal 5% level for corrected p-values. Mediation effects were considered significant if the 95% confidence intervals of indirect effect did not include zero. A detailed reference can be found in the online code.

## 3 Results

### 3.1 Cohort Characteristics

Demographics of the plasma dataset (n=287), the imaging dataset (n=395), and the combined dataset (n=55) are separately presented in Table 1, Supplementary Table 1, and Table 2. Among individuals with Mild Cognitive Impairment (MCI) and Alzheimer’s Disease (AD), we observed a higher likelihood of advanced age in the plasma dataset (p=0.025) and the combined dataset (p=0.071). Additionally, MCI and AD patients exhibited significantly decreased cognitive performance (p<0.0001) in the plasma dataset. When considering plasma biomarkers, significant between-group differences were found for plasma NfL (p<0.001), GFAP (p<0.0001), and p-tau181 (p<0.0001) in both the plasma and combined datasets. However, the plasma Aβ42/Aβ40 was significant only in the plasma dataset (p<0.001), while no significant differences were observed for Aβ42 and Aβ40 (p>0.05). Furthermore, whole-brain generalized local efficiency demonstrated significant differences in both the imaging dataset and the combined dataset (p<0.01).

### 3.2 Plasma markers related to AD progression

Across four diagnostic groups (NC, SCD, MCI and AD), significant varying trends with AD progression for all four plasma markers were observed in the plasma dataset. Specifically, GFAP emerged as the most sensitive marker (linear trend: t=11.164, p=3.59×10^-24^; quadratic trend: t=7.708, p=2.25×10^-13^; adjusted R² = 0.475), followed by NfL (linear trend: t=6.542, p=2.9×10^-10^; quadratic trend: t= 3.896, p=1.22×10^-4^; adjusted R² = 0.330), p-tau181 (linear trend: t= 8.452, p=1.61×10^-15^; quadratic trend: t=6.316, p=1.05×10^-9^; adjusted R² = 0.346) and Aβ42/Aβ40 (linear trend: t=-3.257, p=1.27×10^-3^; quadratic trend: t=-1.662, p=9.76×10^-2^; adjusted R² = 0.101) (Figure 1, Supplementary Table 2). We also observed significant correlations between these plasma markers and general cognitive performances in the plasma dataset (Figure 2, p < .00001 for GFAP, NfL, and p-tau181. See Supplementary Figure 2 for the correlation of all plasma markers and cognition).

**Figure 1:**
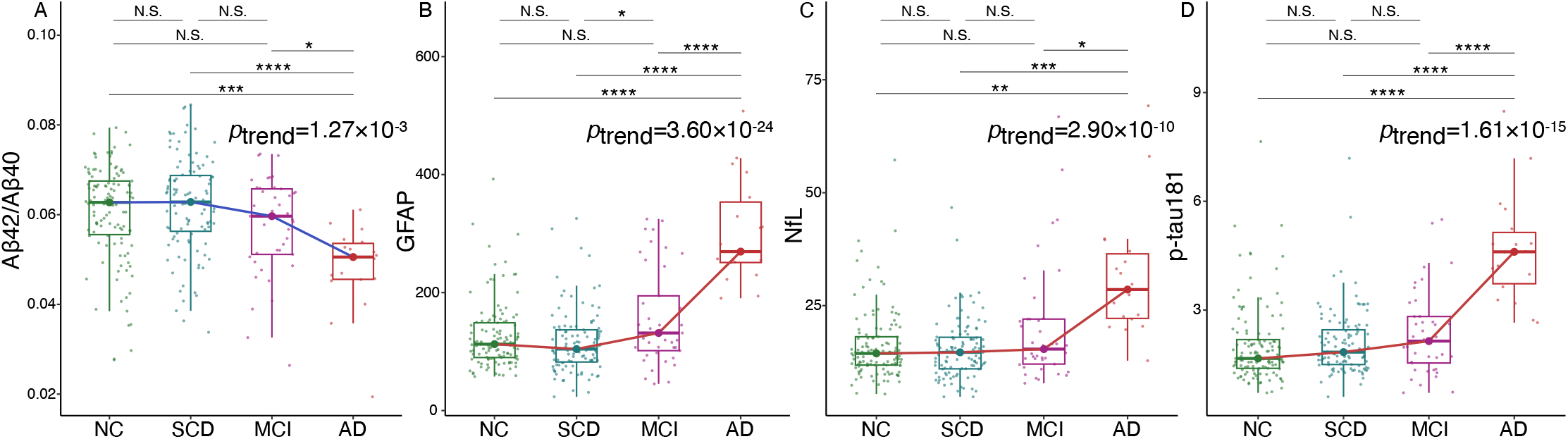
Distribution and Significance of Linear Trends in Plasma Markers of Aβ42/Aβ40 (A), GFAP (B), NfL (C), and p-tau181 (D) among Normal Controls (NC), Subjective Cognitive Decline (SCD), Mild Cognitive Impairment (MCI), and Alzheimer’s Disease (AD) Groups. Aβ, β-amyloid; GFAP, glial fibrillary acidic protein; NfL, Neurofilament light chain; p-tau181, phosphorylated tau181.

**Figure 2:**
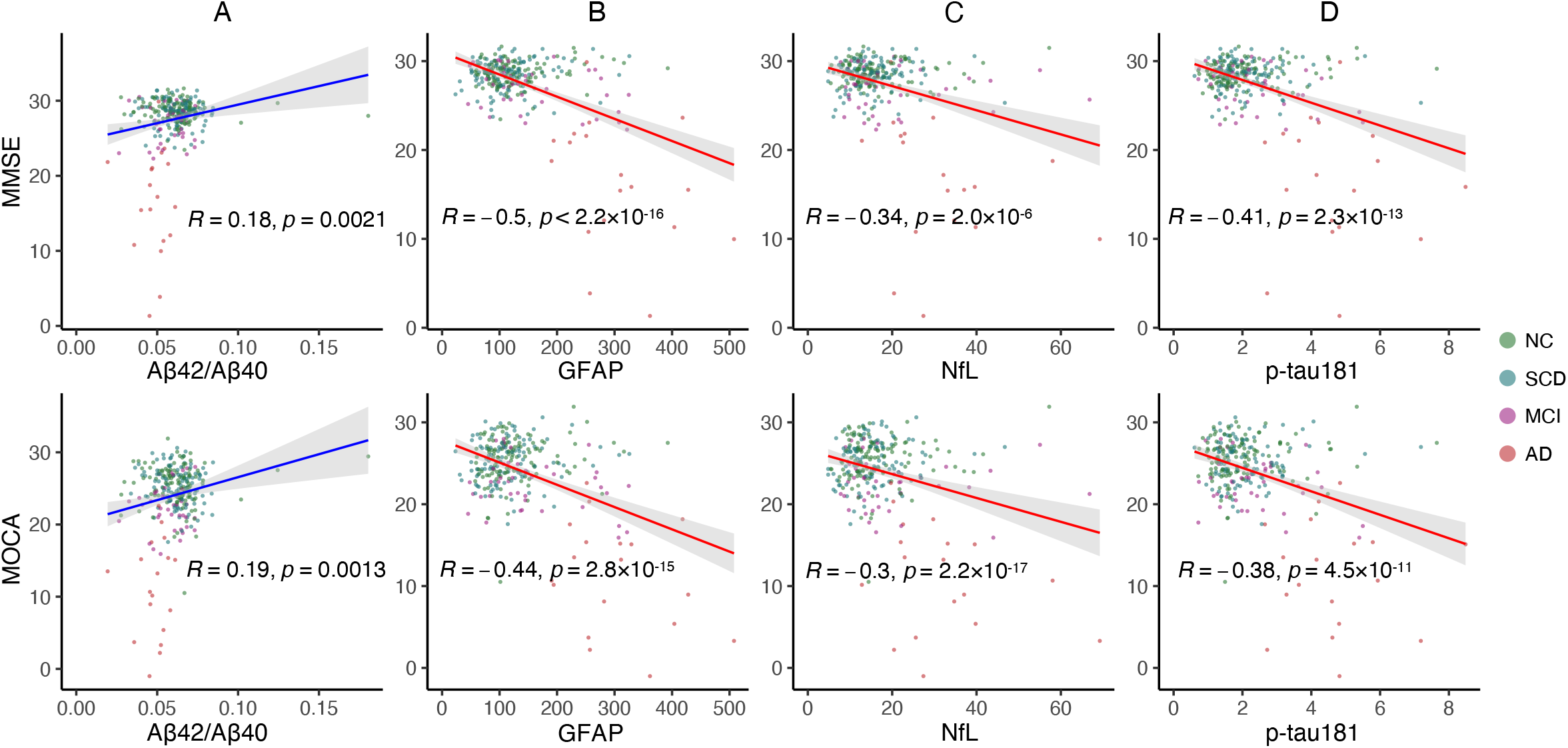
Correlations between Plasma Markers and General Cognition (Upper Panel: MMSE; Lower Panel: MOCA) Controlling for Age, Sex, Years of Education, and APOE4 Status. MMSE, Mini-Mental State Examination; MOCA, Montreal Cognitive Assessment; Aβ, β-amyloid; GFAP, glial fibrillary acidic protein; NfL, Neurofilament light chain; p-tau181, phosphorylated tau181; NC, normal controls; SCD, subjective cognitive decline; MCI, mild cognitive impairment; AD, Alzheimer’s disease.

### 3.3 Decrease network local efficiency with AD progression

Across four diagnostic groups, we found a significant decrease trend in generalized local efficiency of brain structural network in the imaging dataset (linear trend: t=-2.25, p=0.025; quadratic trend: t=-2.58, p=0.011). Further analysis on regional level revealed that 16 brain regions exhibited significant linear decrease (p < 0.05, corrected), mainly located in the bilateral frontal, temporal, parietal cortex and subcortical regions (Figure 3A). Global efficiency and local efficiency didn’t show significant trends at either the whole-brain (Supplementary Table 3) or regional level (Supplementary Table 4). We defined the 16 brain regions with a significant linear trend (p < 0.05, corrected) as AD-Progression-related Regions (ADPRs). Then we extracted the first principal component of generalized local efficiency across ADPRs with the *principal* function from the *psych* package [35] (Figure 3C). The number of components was set to 1 suggested by a prior parallel analysis with 10,000 iterations. As higher local efficiency relates to greater robustness to pathological changes (Figure 3B), the small number of components suggested a highly co-varying pattern among these regions, which may exhibit similar pathological changes across the AD continuum.

**Figure 3:**
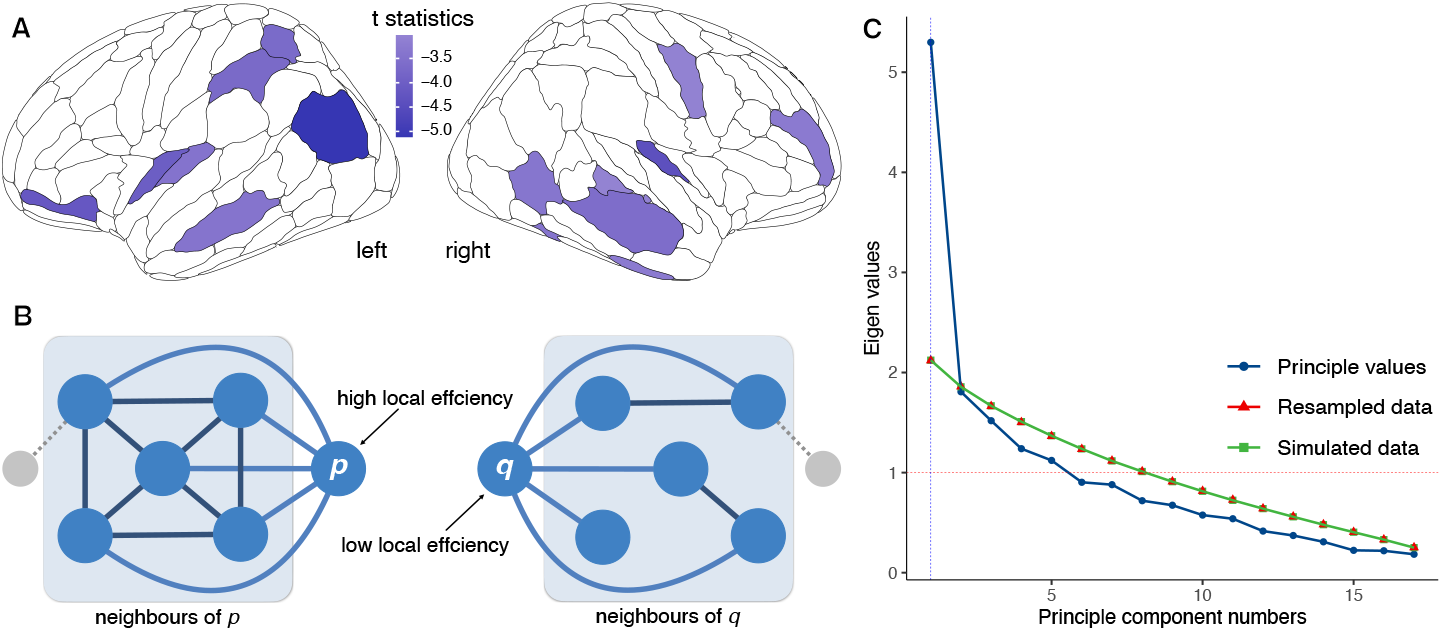
Local Efficiency Disruptions and Principal Component Analysis. (A) Brain regions exhibited a significant linear decrease trend of generalized local efficiency across the AD continuum (p < 0.05, FDR corrected). These regions were mainly located in the bilateral frontal, temporal, parietal cortex, and subcortical regions. (B) Higher local efficiency is associated with greater robustness to pathological changes.(C) Parallel analysis with 10,000 iterations suggested that the number of principal components was one. AD, Alzheimer’s disease; FDR, False Discovery Rate.

### 3.4 Network disruption is related to cognitive decline and plasma markers

We further examined the correlation between network efficiency with plasma markers and cognitive performance in the combined dataset. The component score of generalized local efficiency among ADPRs demonstrated significant correlations with cognitive performance (Pearson’s *R* = 0.66, *p* = 5.7×10^−8^ for MMSE; R = 0.56, p = 9.5×10^−6^ for MOCA) and plasma markers sensitive to AD progression, including GFAP (Pearson’s R = − 0.61, *p* = 6.3×10^−7^), NfL (R = − 0.57, *p* = 6.4×10^−6^), and p-tau181(R = − 0.48, *p* = 2.0×10^−4^). However, no significant correlation was observed with Aβ42/Aβ40 (R = − 0.068, p = 0.62). Notably, the component score of ADPRs exhibited a stronger correlation with pathological and clinical measures (Figure 4A) compared to the whole-brain averaged local efficiency (Figure 4B). The discrepancy may suggest the potential of ADPRs in predicting AD-related changes in plasma markers and cognitive decline. See Supplementary Figure 3 for all the correlations between network efficiency measures and plasma markers as well as cognitive performances.

**Figure 4:**
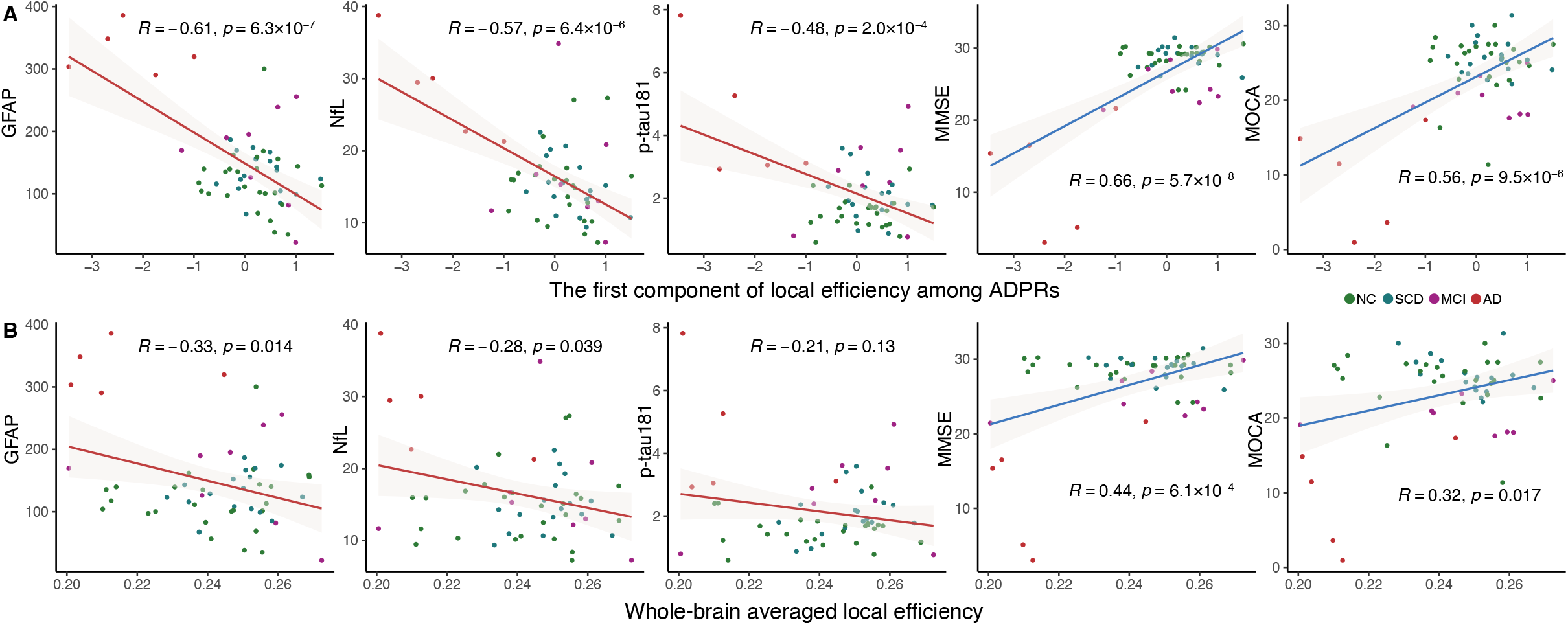
Relationship between Local Efficiency and Plasma Markers with Cognition.

The correlations of (A) the component of generalized local efficiency among AD-Progression-related Regions (ADPRs) were stronger than the correlations with (B) whole-brain averaged efficiency. These correlations were controlled for age, sex, years of dducation, and APOE4 Status. MMSE, Mini-Mental State Examination; MOCA, Montreal Cognitive Assessment; Aβ, β-amyloid; GFAP, glial fibrillary acidic protein; NfL, Neurofilament light chain; p-tau181, phosphorylated tau181; NC, normal controls; SCD, subjective cognitive decline; MCI, mild cognitive impairment; AD, Alzheimer’s disease.

### 3.5 Mediation analysis

To investigate the mediating role of network efficiency in the relationship between plasma markers and cognitive performance, we conducted mediation analyses using both the ADPR component and whole-brain average local efficiency. We found a significant average causal mediation effect, determined through 10,000 bootstrapping, in the relationship between GFAP (ab=-0.224, 95%CI[-0.417, -0.029], p = 0.0196 for MMSE; ab=-0.198, 95%CI[-0.42, -0.003], p =0.0438 for MOCA) and NfL (ab=-0.346, 95% CI [-0.710, -0.036], p=0.0188 for MMSE; ab=-0.300, 95% CI [-0.634, -0.017], p=0.0266 for MOCA) with cognitive performance (Figure 5). The direct effect of NfL on cognitive performance (β=-0.283, p=0.021 for MMSE; β=-0.195, p=0.107 for MOCA) was weaker than the indirect effects mediated by the ADPR score. Notably, we did not observe significant mediation effects through whole brain averaged local efficiency on the relationship between any plasma marker and cognition (Supplementary Table 5).

**Figure 5:**
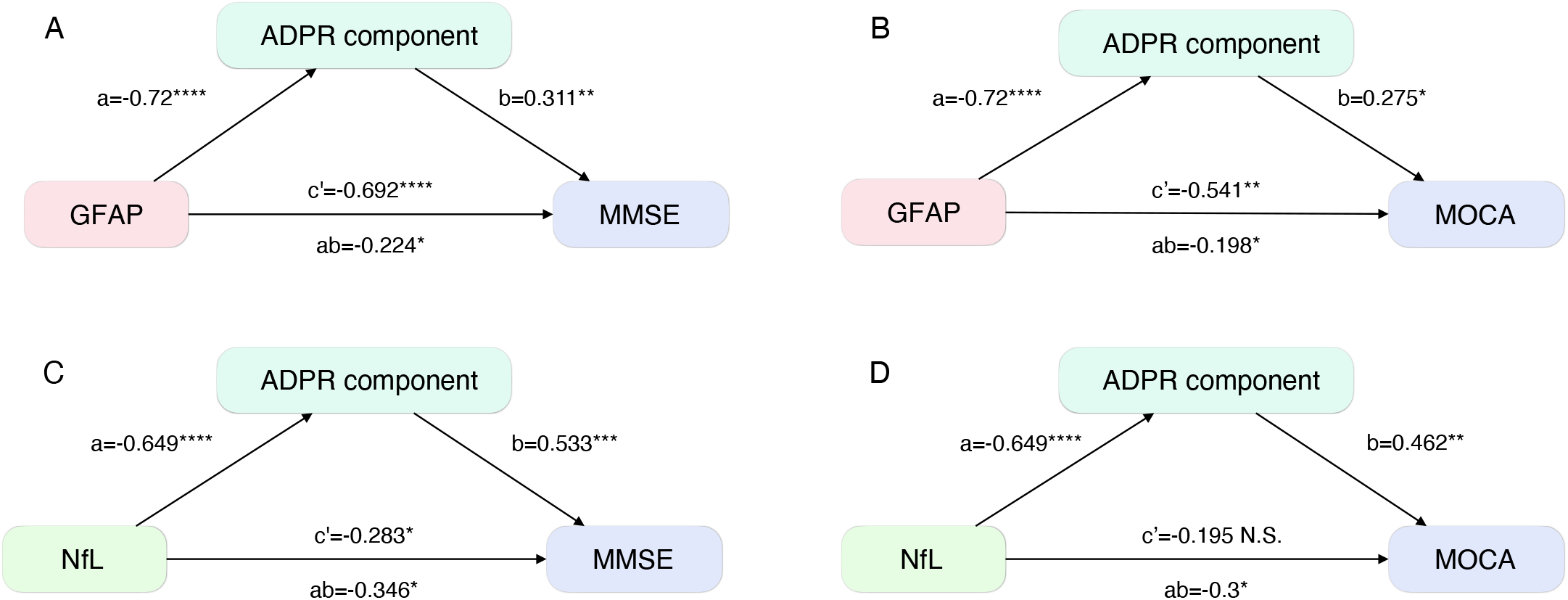
Local Efficiency Mediates the Relationship between Plasma Markers and Cognition. GFAP-MMSE(A), GFAP-MOCA(B), NfL-MMSE(C), NfL-MOCA(D). This mediation effect was found in the AD-Progression-related Regions (ADPRs), determined through 10,000 bootstrapping. MMSE, Mini-Mental State Examination; MOCA, Montreal Cognitive Assessment; GFAP, glial fibrillary acidic protein; NfL, Neurofilament light chain.

## Discussion

Our investigation identified plasma markers and network efficiency highly sensitive to AD progression and cognitive decline. Plasma GFAP emerged as the most sensitive AD progression indicator, strongly correlating with cognitive status. Generalized local efficiency demonstrated a significant downward trend in multiple regions as AD advanced, whose component inversely correlated with plasma GFAP, NfL, and ptau181, while positively correlating with cognitive level (MMSE, MoCA). Integration of plasma biomarkers and magnetic resonance imaging may offer a minimally invasive, cost-effective approach for AD diagnosis, supported by their impact on cognition through network efficiency mediation.

Four plasma biomarkers showed significant trends in the AD progress, including Aβ42/Aβ40, NfL, pTau181, and GFAP. Notably, GFAP exhibited the highest sensitivity, displaying significant variations at the early stage and increasing levels as the disease advanced[36,37]. The literature consistently supports plasma biomarkers’ diagnostic utility in AD, individually or in combination with other markers[36,38–41]. Elevated plasma GFAP levels were observed in cognitively intact individuals with Aβ positivity, particularly distinguishing Aβ+ from Aβ-individuals[12]. GFAP emerged as the earliest and most significantly altered biomarker from preclinical to symptomatic AD, predicting progression and cognitive decline[11,42]. While GFAP levels can be influenced by other neuroinflammatory factors[37], integrating multiple plasma biomarkers with structural efficiency can enhance the precision of staging patients along the AD continuum. Associations between plasma biomarkers and cognitive changes were evident, with GFAP exhibiting notable sensitivity. Previous studies have also demonstrated a strong correlation between plasma GFAP levels and cognition in AD patients[43,44], further validating its diagnostic value in assessing AD trajectory and predicting progression.

We observed a significant declining trend in whole-brain generalized local efficiency from the cognitively normal stage to SCD, MCI, and AD. Recent studies have shown a discernible decline in local efficiency as AD advances and a noteworthy correlation between network efficiency and hallmark pathological features of AD, like Aβ deposition and tau tangles[18–20]. In conjunction with existing literature, our findings underscore the potential of local efficiency as a non-invasive biomarker to detect AD-related brain pathological changes.

We found regional disruptions in both hub regions (insular and superior parietal lobule) and peripheral regions (inferior parietal lobule, lateral temporal lobe, striatum, orbital gyrus, and middle frontal gyrus). The literature on the topological disruptions along the AD continuum [17,45–47] supports the outcomes of our investigation, highlighting the potential of network efficiency as a cost-effective, effective, and non-invasive biomarker for AD imaging. Moreover, we note that the distribution of ADPRs overlaps with the pathological progression frequently observed in frontal, cingulate, precuneus, striatum, parietal, and lateral temporal cortices[14,48]. The cerebral regions identified in our study closely align with confirmed sites of AD pathology deposition, strongly indicating that pathological deposition may contribute to deviations in local efficiency, ultimately resulting in cognitive decline. Taken together, We propose that local efficiency may capture a broad spectrum of brain damage across different AD stages.

The mechanisms underlying the impact of pathological molecules on cognitive decline along the AD continuum are complex. Building upon the possible mediation effect of neuroimaging biomarkers on the link between cognitive function and plasma biomarkers, our study delved into their intricate connections, specifically focusing on GFAP and NfL. Importantly, we found that the principal component of local efficiency among AD pathological regions exhibited robust correlations with plasma biomarkers and cognitive function, surpassing the overall brain local efficiency. Notably, these mediation effects were only significant within the principal components among ADPRs, rather than the average efficiency of the entire brain. This observation aligns with previous studies revealing significant reductions in local efficiency within the AD population[49]. Moreover, individuals carrying the apolipoprotein E ε4 allele displayed even greater reductions in local efficiency, underscoring its role as a reliable and distinctive indicator across the AD continuum[50]. Overall, our study contributes novel insights into the complex mechanisms driving the pathological cognitive decline in the AD process, elucidating the pivotal role of network efficiency.

Several limitations should be considered in interpreting the findings of this study. Firstly, it is important to note that this preliminary investigation was conducted at a single center with a relatively modest sample size. Future studies should involve larger sample sizes from multiple centers to enhance the robustness and generalizability of our results. Secondly, it is worth mentioning that not all participants in the SILCODE cohort had complete blood and imaging data, leading to the division of the cohort into separate datasets for analysis. Collecting comprehensive clinical data during recruitment would facilitate more thorough follow-up investigations. Thirdly, it is noteworthy that amyloid-PET data were unavailable for all individuals with Alzheimer’s disease and mild cognitive impairment included in our study. Consequently, the diagnostic process relied primarily on clinical data and imaging markers[51,52]. Future research should incorporate ample gold-standard evidence, including amyloid-PET imaging, to strengthen diagnostic validity. Finally, due to the longitudinal nature of the SILCODE cohort, all subjects were initially free from cognitive impairment, resulting in potential variations in age and gender distribution within our sample. The limited number of cases identified during the investigation of mild cognitive impairment and Alzheimer’s disease may be attributed to this baseline characteristic. Extending the follow-up duration would allow for more substantial findings. Adhering to these considerations in future studies will enhance the validity and clinical applicability of our findings in the field of neurology.

## Conclusions

In conclusion, our study highlights the significance of plasma GFAP levels as a valuable indicator for the early detection and prognosis of AD. We also emphasize the utility of generalized local efficiency in relevant brain regions as a sensitive marker for tracking disease progression. Furthermore, our findings demonstrate the significant mediation of network efficiency among AD-related regions in the relationship between plasma markers and general cognition. These results underscore the potential of integrating brain network connectivity efficiency and plasma biomarkers as a cost-effective strategy for screening and diagnosing AD. Combining these approaches holds promise in enhancing the accuracy and efficiency of AD diagnosis and prognosis, ultimately benefiting patients and advancing our understanding of the disease. Further research in larger cohorts is warranted to validate and expand upon these findings.

## Data Availability

All data produced in the present study are available upon reasonable request to the authors

## Consent Statement

All participants or their legal guardians provided written informed consent. All procedures were approved by the Medical Research Ethics Committee and Institutional Review Board of Xuanwu Hospital. Cohorts are registered with ClinicalTrials.gov (SILCODE: NCT02225964).

## Acknowledgment

The authors would like to thank Capital Medical University Xuanwu Hospital and the State Key Laboratory of Cognitive Neuroscience and Learning, Beijing Normal University for assistance with this study. The authors also express their gratitude to the Sino Longitudinal Study on Cognitive Decline (SILCODE) project in China for funding this study’s data collection and sharing aspects.

We thank the authors for their contributions as follows. Mingkai Zhang, Haojie Chen: Investigation, Data curation, Writing - original draft, Writing - review & editing. Weijie Huang: Investigation, Writing - original draft. Tengfei Guo: Data curation, Writing – review. Guolin Ma: Writing – review. Ying Han, Ni Shu: Investigation, Data curation, Writing - original draft, Writing - review & editing, Supervision.

## Source of Funding

This study was supported by the Sino-German Cooperation Grant (M-0759); STI2030-Major Projects (2022ZD0213300, 2022ZD0211800); National Natural Science Foundation of China (82020108013, 82001773, 32271145, 81871425); Fundamental Research Funds for the Central Universities (2017XTCX04); Open Research Fund of the State Key Laboratory of Cognitive Neuroscience and Learning (CNLZD2101, CNLYB2001).

## Disclosures

None

## Notes

### Competing Interest Statement

The authors have declared no competing interest.

