## Supplementary Tables for "Relationship between topological efficiency of white matter structural connectome and plasma biomarkers across Alzheimer’s disease continuum"

|  | NC(n=121) | SCD(n=107) | MCI(n=41) | AD(n=18) | Overall(n=287) | Statistics(P) |
| --- | --- | --- | --- | --- | --- | --- |
| Sex(M/F) | 42/79 | 33/74 | 19/22 | 7/11 | 101/186 | 3.243(0.518) |
| Age | 69.0±5.58 | 68.1±7.28 | 69.8±8.02 | 73.8±6.81 | 69.1±6.80 | 2.803(0.025) |
| Education | 12.4±3.17 | 13.2±3.51 | 10.9±4.05 | 12.4±2.66 | 12.5±3.48 | 3.418(<.01) |
| APOEε4 carriers  (percentage) | 18.2% | 34.6% | 41.5% | 66.7% | 30.7% | 22.862(<.001) |
| MMSE | 28.8±1.19 | 28.6±1.52 | 26.5±2.63 | 15.4±6.55 | 27.5±3.93 | 73.054(<.0001) |
| MoCA-B | 25.7±2.89 | 25.6±2.48 | 21.6±3.21 | 9.89±5.94 | 24.1±4.98 | 64.563(<.0001) |
| Plasma Aβ42(pg/mL) | 6.26±1.45 | 6.14±1.53 | 6.22±1.52 | 5.17±1.60 | 6.14±1.51 | 2.111(0.078) |
| Plasma Aβ40(pg/mL) | 104±21.1 | 99.5±21.6 | 108±17.8 | 109±26.9 | 103±21.4 | 1.755(0.137) |
| Plasma Aβ42/Aβ40 | 0.0624±0.0167 | 0.0623±0.0106 | 0.0575±0.0109 | 0.0482±0.0095 | 0.0607±0.0139 | 5.092(<.001) |
| Plasma NfL(pg/mL) | 16.2±7.80 | 15.3±6.66 | 20.0±13.0 | 31.5±14.0 | 17.4±9.64 | 13.073(<.0001) |
| Plasma GFAP(pg/mL) | 126±56.5 | 116±54.0 | 156±82.3 | 303±88.9 | 138±76.4 | 29.455(<.0001) |
| Plasma p-tau181  (pg/mL) | 2.03±1.07 | 2.09±0.91 | 2.35±1.16 | 4.67±1.50 | 2.27±1.23 | 21.764(<.0001) |

Table 1 Sample characteristics in the blood dataset.

Continuous data were presented as mean ± SD. Analyses of variance were utilized for continuous variables, while chi-square tests were employed for categorical variables across NC, SCD, MCI, and AD. Abbreviations: APOE, apolipoprotein E; MMSE, Mini-Mental State Examination; MoCA-B, Montreal cognitive assessment basic; Aβ, amyloid β; NfL, neurofilament light chain; GFAP, glial fibrillary acidic protein; p-tau, phosphorylated tau; NC, normal controls; SCD, subjective cognitive decline; MCI, mild cognitive impairment; AD, Alzheimer's disease.

|  | NC(n=25) | SCD(n=17) | MCI(n=8) | AD(n=5) | Overall(n=55) | Statistics(P) |
| --- | --- | --- | --- | --- | --- | --- |
| Sex(M/F) | 8/17 | 5/12 | 3/5 | 1/4 | 17/38 | 0.473(0.976) |
| Age | 65.7±4.98 | 61.4±8.03 | 69.5±9.55 | 69.2±8.91 | 65.2±7.53 | 2.228(0.071) |
| Education | 12.6±3.21 | 13.5±3.94 | 13.1±3.52 | 13.0±4.24 | 13.0±3.50 | 0.162(0.957) |
| APOEε4 carriers  (percentage) | 28.0% | 35.3% | 75.0% | 40.0% | 38.2% | 5.76(0.218) |
| MMSE | 28.8±1.48 | 29.0±1.17 | 24.8±3.11 | 12.0±7.91 | 26.7±5.61 | 16.406(<.0001) |
| MoCA-B | 24.9±3.88 | 26.5±2.45 | 19.4±2.83 | 8.80±5.76 | 23.1±6.17 | 13.688(<.0001) |
| Plasma Aβ42(pg/mL) | 6.09±1.21 | 5.71±1.85 | 5.98±1.27 | 4.84±1.48 | 5.84±1.47 | 0.808(0.523) |
| Plasma Aβ40(pg/mL) | 100±20.2 | 93.5±28.3 | 111±15.5 | 96.9±37.3 | 99.4±24.2 | 0.754(0.558) |
| Plasma Aβ42/Aβ40 | 0.0631±0.0155 | 0.0625±0.0115 | 0.0540±0.0102 | 0.0517±0.00658 | 0.0605±0.0135 | 1.358(0.253) |
| Plasma NfL(pg/mL) | 15.1±6.05 | 13.1±4.82 | 18.8±8.88 | 30.8±7.68 | 16.4±7.89 | 6.429(<.001) |
| Plasma GFAP(pg/mL) | 114±47.6 | 113±49.9 | 201±105 | 359±69.9 | 149±94.7 | 11.43(<.0001) |
| Plasma p-tau181  (pg/mL) | 1.53±0.526 | 1.88±0.729 | 3.07±1.32 | 4.66±2.28 | 2.14±1.34 | 8.887(<.0001) |
| Global efficiency | 6.55±5.71 | 5.18±2.94 | 5.05±3.49 | 5.87±6.30 | 5.85±4.69 | 0.278(0.891) |
| Local efficiency | 13.2±13.4 | 10.5±6.23 | 10.5±7.73 | 12.3±11.1 | 11.9±10.5 | 0.21(0.932) |
| Generalized local efficiency | 0.242±0.0175 | 0.249±0.0112 | 0.245±0.0211 | 0.213±0.0167 | 0.242±0.0186 | 4.182(<.01) |

TABLE 2 Sample characteristics in the combined dataset with network efficiency.

Continuous data were presented as mean ± SD. Analyses of variance were utilized for continuous variables, while chi-square tests were employed for categorical variables across NC, SCD, MCI, and AD. Abbreviations: APOE, apolipoprotein E; MMSE, Mini-Mental State Examination; MoCA-B, Montreal cognitive assessment basic; Aβ, amyloid β; NfL, neurofilament light chain; GFAP, glial fibrillary acidic protein; p-tau, phosphorylated tau; NC, normal controls; SCD, subjective cognitive decline; MCI, mild cognitive impairment; AD, Alzheimer's disease.
