## Supplementary FIgures for "Relationship between topological efficiency of white matter structural connectome and plasma biomarkers across Alzheimer’s disease continuum"

### Supplementary Figure 1

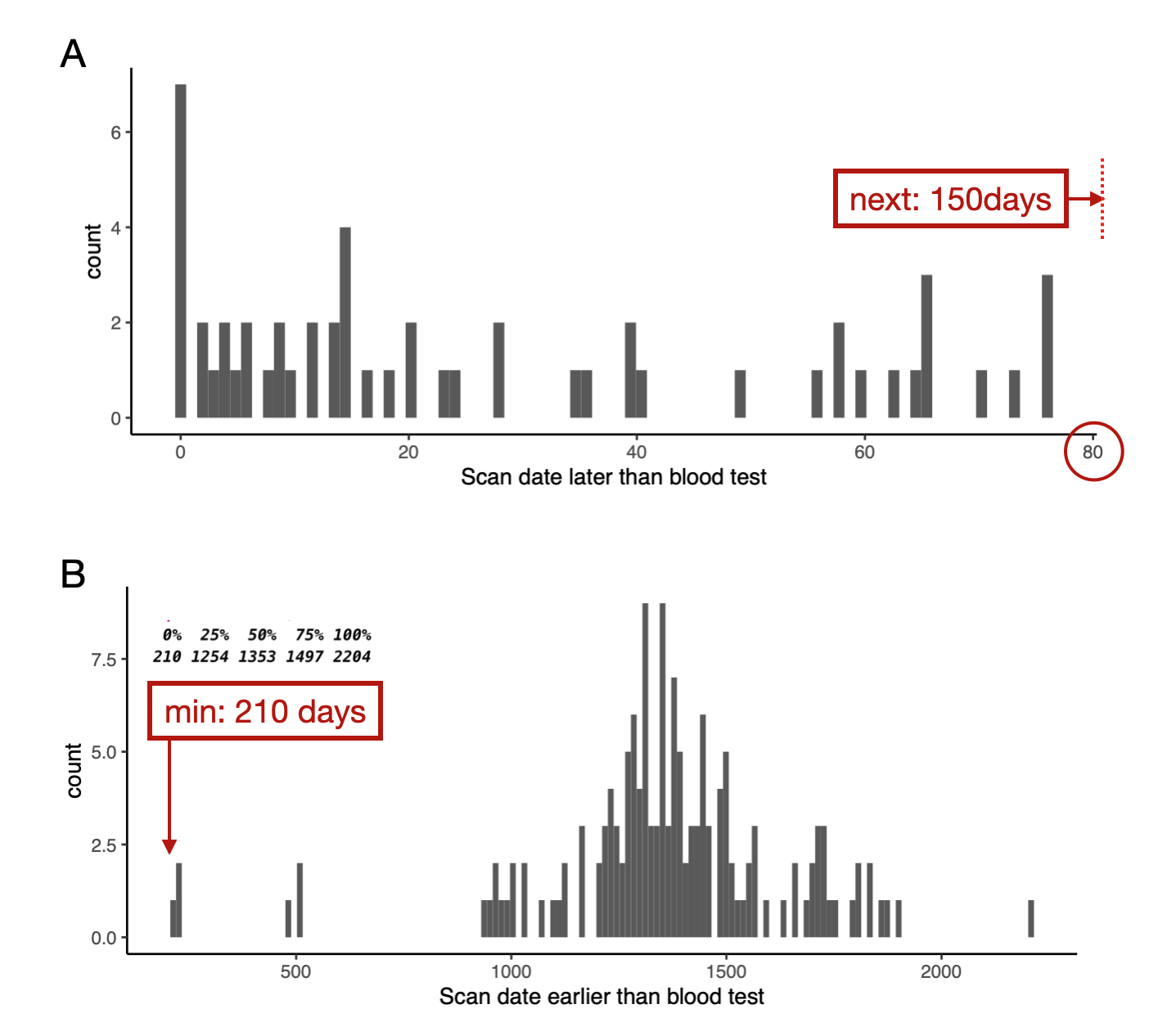

Determining the matching interval between the plasma dataset and imaging dataset to create the combined dataset. Participants in the first two datasets who matched plasma and imaging time differences of not more than 80 days.

### Supplementary Figure 2

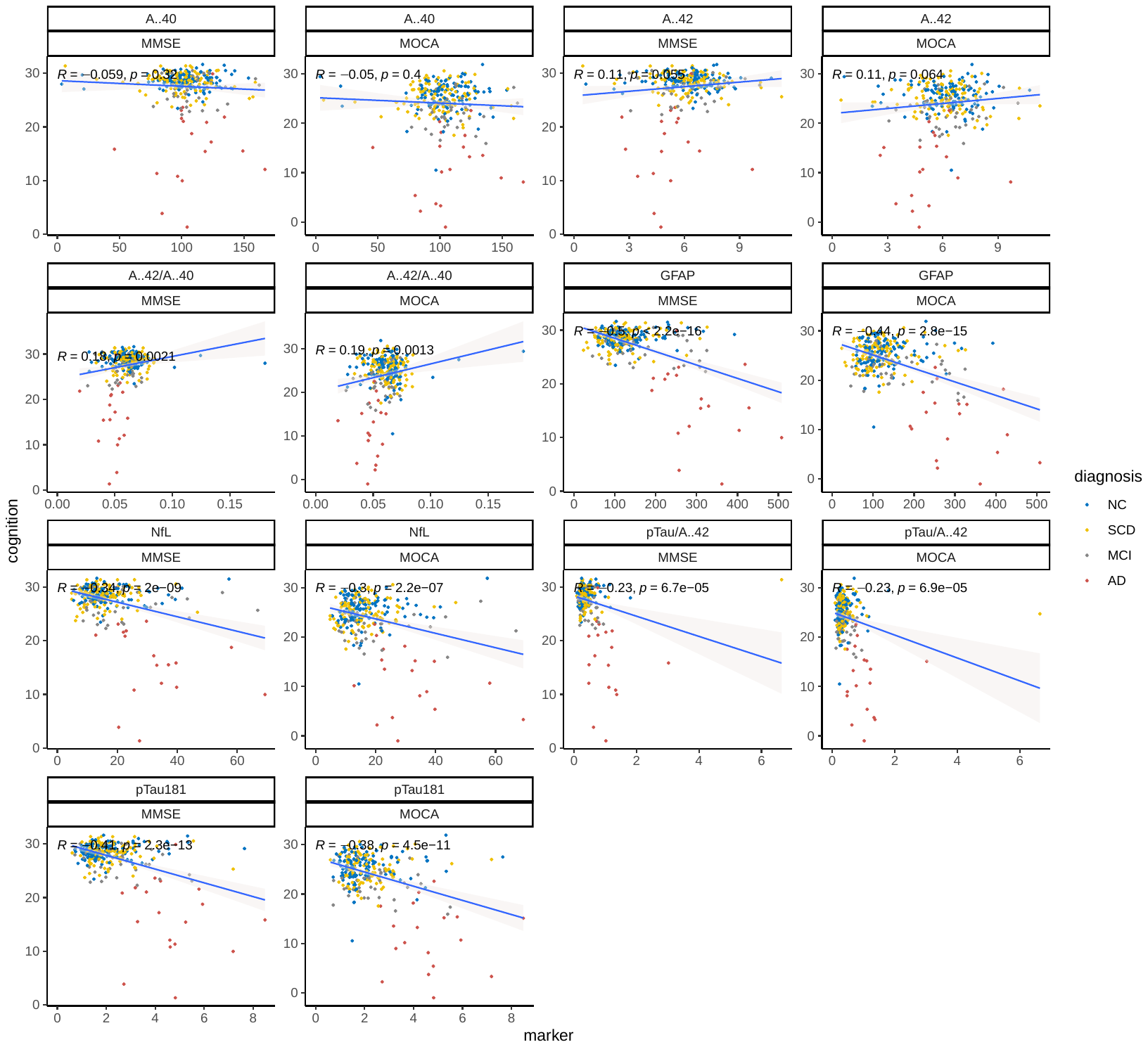

**Partial correlation of plasma markers and general cognitions, controlling for age, sex, years of education, and APOE4 status.**

### Supplementary Figure 3

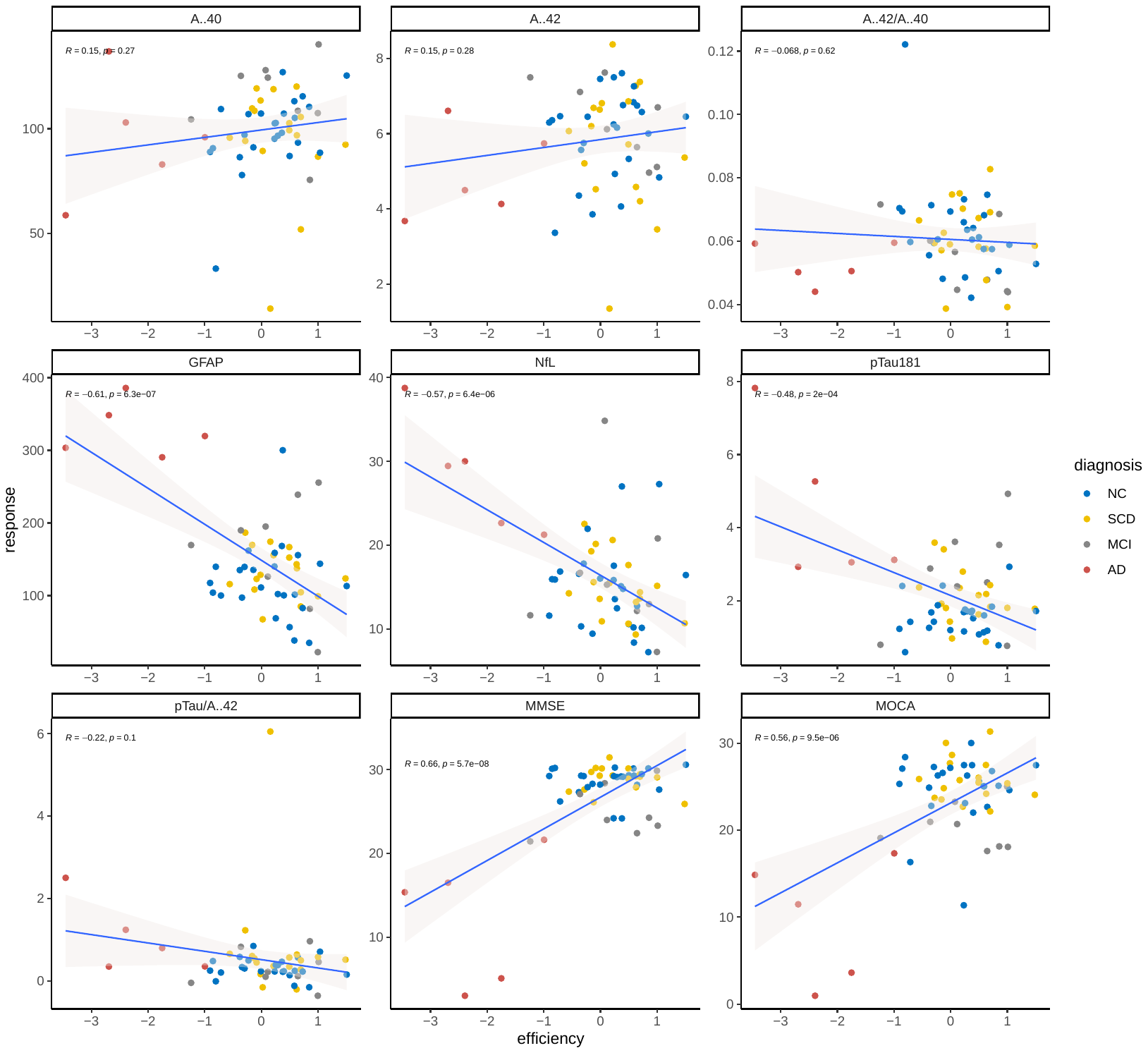

**Partial correlation of the component of generalized local efficiency among ADPRs and bio-measures, controlling for age, sex, years of education, and APOE4 status.**
